# Validity of observation tools for patients hospitalized with osteoporotic vertebral fractures

**DOI:** 10.1101/2023.06.02.23290859

**Authors:** Youhei Yoshimi, Takanori Matsuura, Kazuaki Miyazato, Shiho Takahashi, Nami Tanaka, Hanae Morinaga, Asuka Hayata, Minami Onishi, Yousuke Nagano, Hideo Ohnishi

## Abstract

Osteoporotic vertebral fractures in older patients impair activities of daily living because of low back pain and abnormal posture. Assessing pain using self-reported assessment tools is difficult, especially in patients with moderate-to-severe cognitive impairment. In recent years, observational assessment tools have been used when self-reported assessment tools were difficult to implement. However, no reports have investigated the usefulness of observational assessment tools in patients with acute-phase orthopedic disorders without comorbidities. This study examined the validity of observational tools for pain assessment in patients with lumbar vertebral fractures. Patients admitted to our hospital with acute-phase vertebral fractures were enrolled in this prospective study. Pain was assessed using Japanese versions of the Abbey Pain Scale and Doloplus-2 observational assessment tools, in addition to the Numerical Rating Scale, a self-reported assessment tool. To verify the validity of each pain assessment tool, we examined whether each tool correlated with the activities of daily living and ambulatory status. Activities of daily livings were assessed using the Barthel Index. Ambulatory status was assessed using the Functional Ambulation Categories and the 10-m walking test. Similar to the Numerical Rating Scale scores, assessments with the Abbey Pain Scale and Doloplus-2 showed significant decreases in scores over time. In addition, a significant positive correlation was observed between the self-reported and observational assessment tools. Each pain assessment tool was significantly negatively correlated with activities of daily livings and ambulatory status. Our results indicated when self-reported assessment with the Numerical Rating Scale was difficult for patients with cognitive impairment, pain could be estimated using the Abbey Pain Scale and Doloplus-2 observational assessment tools.

## Introduction

Osteoporotic vertebral fractures occur in older adults and result in a functional loss in activities of daily living (ADLs) because of prolonged lower back pain and abnormal posture [1–5]. In Japan, recent reports have that shown musculoskeletal disorders are one of the main reasons older adults require nursing care due to falls and resultant fractures [6]. Thus, preventing severe osteoporosis and low-trauma fractures is also important in terms of health economics [2, 7]. The annual incidence of vertebral fractures in Japan has been reported to be approximately 1,558 per 100,000 people (females 2,117, males 729; the ratio of women to men was 2.90; Kure City, Hiroshima Prefecture, 2015 population aging rate: 32.9%) [8].

Conservative orthotic treatment is the major treatment approach for fresh osteoporotic vertebral fractures, except for burst fractures [9, 10]. At our hospital, patients who have difficulty receiving outpatient treatment are hospitalized for treatment aimed primarily at pain control using bracing medication and rehabilitation to prevent muscle atrophy. The length of hospital stay may be long depending on the pathological condition, physical activity, and social background of the patient, such as living alone.

A recent meta-analysis have shown that pain cannot fully explain the decrease in quality of life in patients with osteoporotic fractures [3]. Pain is difficult to assess in cognitively impaired patients with osteoporotic vertebral fractures, and the validity of pain assessments may be questionable [11–16]. The International Association for the Study of Pain recommends verbal communication tools such as self-report assessments for patients with early cognitive memory decline [4]. However, these tools are difficult to use when assessing patients with advanced dementia [17, 18]. In recent years, the use of observational assessment tools has been proposed as an alternative approach for patients in instances where self-reported pain assessment is difficult to perform [19–24]. However, no studies have been reported on the usefulness of observational assessment tools for patients with acute-phase orthopedic disorders without comorbidities, and it is unknown which observational assessment tool is the most appropriate to use [25].

Moji Ward in Kitakyushu City is one of the leading super-aging districts in Japan. In March 2020, it was estimated that 36.5% of the population was aged 65 years or older (94,355 individuals; male-female composition: 43,205 males and 51,150 females). Many of these patients who have been hospitalized experience cognitive decline. Optimizing pain assessment tools for patients with cognitive decline is an important issue in acute medical care for providing adequate pain relief and rehabilitation of osteoporotic vertebral fractures [11, 13–15].

This study aimed to prospectively evaluate the validity of observational assessment tools. Pain in patients who were admitted to our hospital with acute-phase vertebral fractures was assessed using the Japanese version of the Abbey Pain Scale (Abbey-J) [26] and Doloplus-2 [27], which are observational assessment tools, in addition to administering the Numerical Rating Scale (NRS), a self-reported assessment tool. The scores of each assessment tool were examined for changes and correlations over time after admission. Furthermore, to evaluate their validity, we determined whether each pain assessment tool correlated with ADLs and ambulatory status.

## Materials and methods

### Participants and ethical approval

Thirty-five patients aged 65 years or older who visited the Moji Medical Center with the chief complaint of low back pain before subsequently being hospitalized and diagnosed with a vertebral body fracture between April 2022 and March 2023 were prospectively enrolled. Exclusion criteria included: (1) comorbidities; (2) requirements to stay in bed; or (3) inability to participate in the rehabilitation programs. All patients received an explanation of the study and provided their written consent to participate on admission.

This study was approved by the ethical review board of the Moji Medical Center (approval no: 02-01) and was conducted in compliance with the Declaration of Helsinki. We disclosed this information to the subjects and provided them with the opportunity to refuse consent.

### Assessment

#### Pain assessments using self-reported and observational assessment tools

Nurses assessed the pain of the patients at rest and during movement (during transfer to the bathroom) over 10 consecutive days from the day of admission using the NRS (a self-reported assessment tool) and Abbey-J (an observational assessment tool). The NRS is a verbal communication tool in which patients rate pain on an 11-point scale ranging from 0 (no pain) to 10 (worst pain). For the Abbey-J, pain is rated on a scale of 0 to 3 points (a maximum total score of 18 points) on six items reflecting behaviors such as changes during specified movements, vocalizations, and facial expressions. The pain intensity was graded into 4 grades: no pain (0–2 points), mild pain (3–7 points), moderate pain (8–13 points), or severe pain (14–18 points).

In addition, information on the living conditions of each patient was surveyed, and nurses assessed their pain twice a week (days 4 and 7) with the Doloplus-2, an observational tool developed for older people with chronic pain who are unable to complain of pain to others. The pain intensity was scored from 0 to 3, with higher scores indicating more severe pain. The maximum score is 30 points, with a score of 5 points or higher indicating the presence of pain.

The assessments were performed by the nurses in charge of the patients on the corresponding days, and the evaluators were not fixed. In consideration of the variability among evaluators, the mean scores on days 1–4, 5–7, and 8–10 were calculated for analyses.

#### Assessment of ambulatory status and ADLs

On admission, the attending physician checked the patient’s ambulatory status before the injury through history-taking. After the patients were instructed to wear a corset, the physical therapist also evaluated the ADLs using the Barthel Index (BI) once a week during the hospital stay, and ambulatory status using the Functional Ambulation Categories (FAC) and 10-m walking test once a week (days 7, 14, 21, 28, and 35). The BI is an assessment tool for ADLs which is scored out of 100 points on all 10 items, according to the classification of independence, partial assistance, and total dependence. The 10 items include eating, moving, grooming, toileting, bathing, walking (moving), going up/downstairs, dressing, defecating, and urinating. The FAC, originally used in patients with stroke, is a clinical assessment index of walking ability based on the amount of assistance. Walking ability was classified into six categories (score 0: nonfunctional ambulator, score 1: ambulator, dependent on physical assistance – level I, sore 2: ambulator, dependent on physical assistance – level II, score 3: ambulator, dependent on supervision, score 4: ambulator, independent level surface only, score 5: ambulator, independent based on the observation of movement.

#### Evaluation of pain assessment tool scores using the Mini-Mental State Examination (MMSE)

The MMSE is a neuropsychological screening test for dementia used to objectively determine which cognitive function is impaired and to what extent (out of 30 points, a score of ≤ 23 points strongly suggests cognitive decline) [28]. Additionally, MMSE-J is the authorized Japanese translation published by Nihon Bunka Kagaku sha under the permission of Psychological Assessment Resources, Inc. Individuals who purchased the MMSE-J Test Forms provided permission to use them as part of the research. MMSE scores classify the degrees of cognitive decline as normal (score 24–30), mild (score 18–23), or moderate to severe (score 0–17).

### Data analysis

A univariate analysis was performed to compare the groups using repeated analysis of variance (ANOVA) and one-way ANOVA. Considering multicollinearity, Pearson’s correlation coefficient was calculated to describe the associations between the evaluation items. A multivariate logistic regression analysis was performed after analyzing the Pearson’s correlation coefficient.

## Results

### Participant characteristics

Thirty-five patients (mean age 84.4±6.65 years, range 67-95 years; 31 females) with osteoporotic vertebral fractures were included. The average height and weight were 147.3±9.46 cm (123-163 cm) and 46.3±9.46 kg (31.3-76.0 kg), respectively. The site and incidence of each fracture was Th7 (1 patient), Th8 (2 patients), Th9 (1 patient), Th10 (2 patients), Th11 (2 patients), Th12 (12 patients), L1 (6 patients), L2 (7 patients), L3 (3 patients), L5 (1) (Table 1).

**Table 1.** Patient characteristics.

|  |  |
| --- | --- |
| Age | 84.4 ±6.65 |
| Sex; male : female | 4(11.4%): 31 (88.6%) |
| Height (cm) | 147.3 ±7.97 |
| Weight (kg) | 46.3 ±9.46 |
| Body mass index | 21.3 ±4.02 |
| Fracture (number)<br>(multiple fracture included) | Th7 ( 1 ), Th8 ( 2 ), Th9 ( 1 ), Th10 ( 2 ),<br>Th11 ( 2 ), Th12 ( 12 ), L1 ( 6 ), L2 ( 7 ),<br>L3 ( 3 ), L5 ( 1 ) |

### Changes in the scores on each pain assessment tool over time

The NRS, Abbey-J, and Doloplus-2 scores decreased significantly over time after admission (repeated-measures ANOVA followed by the Bonferroni test; NRS at rest): F7, 140 = 9.5035, p = 0.000000001263; at 0.5 week vs. at 1.0 weeks: p = 0.0409, vs. at 1.5 weeks: p = 0.0021, vs. at 2.0 weeks: p = 0.0032, vs. at 2.5 weeks: p = 0.0029, vs. at 3.0 weeks: p = 0.0248, vs. at 3.5 weeks: p = 0.0024, vs. at 4.0 weeks: p = 0.0029: NRS (during movement): F7,133 = 22.243, p <0 .0001; at 0.5 week vs. at 1.0 weeks, p <0 .0001, vs. at 1.5 weeks: p <0 .0001, vs. at 2.0 weeks: p <0 .0001, vs. at 2.5 weeks: p <0 .0001, vs. at 3.0 weeks: p <0 .0001, vs. at 3.5 weeks: p <0 .0001, vs. at 4.0 weeks: p <0 .0001: Abbey-J (at rest): F7,133 = 3.012, p = 0.005718; at 0.5 week vs. at 1.0 weeks: p = 1, vs. at 1.5 weeks: p = 1, vs. at 2.0 weeks: p = 1, vs. at 2.5 weeks: p = 1, vs. at 3.0 weeks: p = 1, vs. at 3.5 weeks: p = 1, vs. at 4.0 weeks: p = 1; Abbey-J (during movement): F7,133 = 3.9269, p = 0.0006321; at 0.5 week vs. at 1.0 weeks: p = 0.217, vs. at 1.5 weeks: p = 1, vs. at 2.0 weeks: p = 1, vs. at 2.5 weeks: p = 0.024, vs. at 3.0 weeks: p = 0.099, vs. at 3.5 weeks: p = 0.458, vs. at 4.0 weeks: p = 0.259; Doloplus-2: F7, 133 = 11.014, p <0 .0001; at 0.5 week vs. at 1.0 weeks: p = 0.00643, vs. at 1.5 weeks: p <0 .0001, vs. at 2.0 weeks: p <0 .0001, vs. at 2.5 weeks: p = 0.00032, vs. at 3.0 weeks: p = 0.0046, vs. at 3.5 weeks: p = 0.0006, vs. at 4.0 weeks: p = 0.00367 (Fig 1).

**Fig 1.**
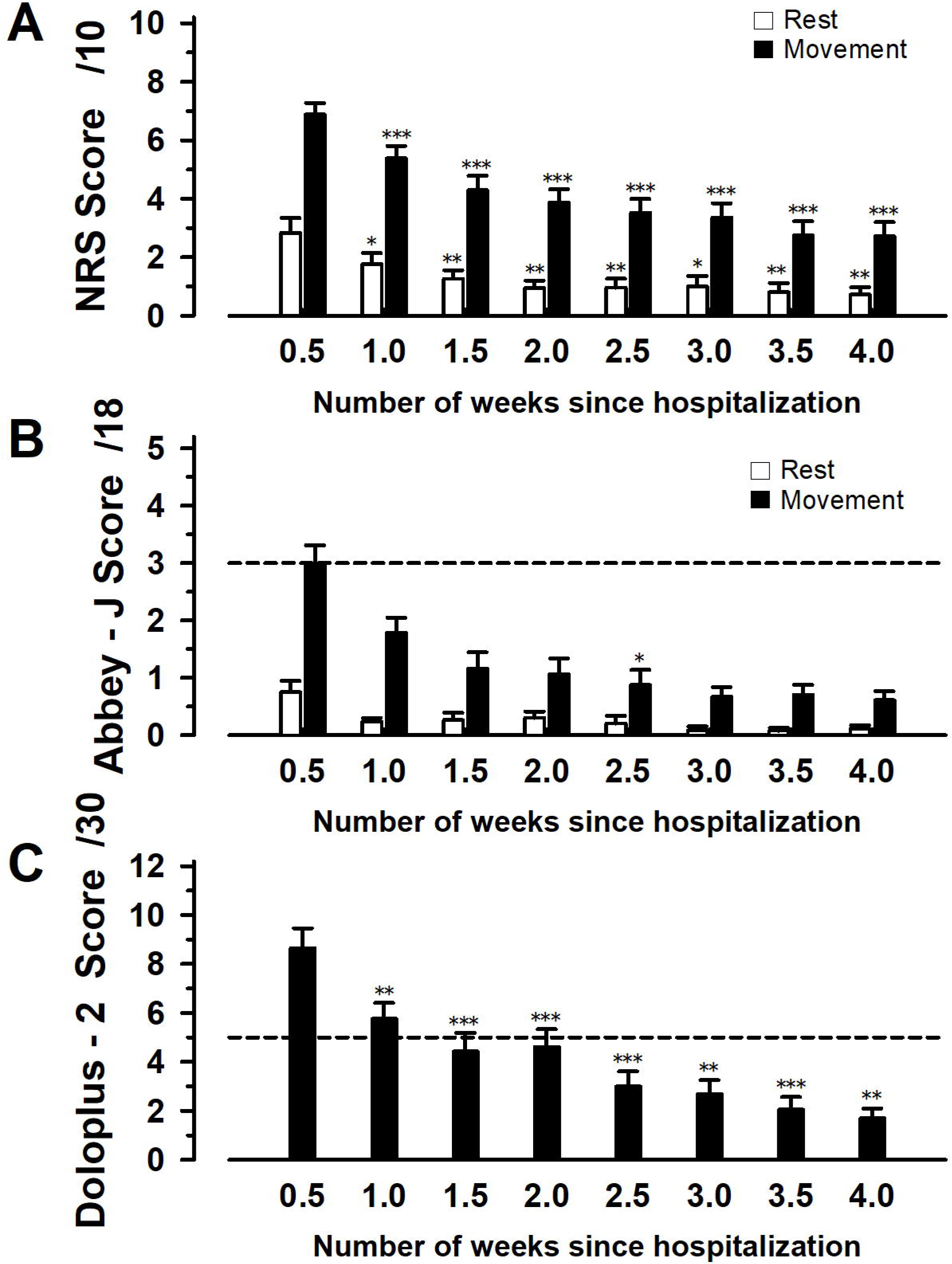
Changes in scores for each pain assessment tool in hospitalized patients with vertebral fractures. (A) Numerical Rating Scale (NRS); (B) Japanese version of Abbey Pain Scale (Abbey-J); (C) Doloplus-2. **p value < 0.01 and ***p value < 0.01 are statistically significant.

Next, we evaluated the rate of change in scores over the course of hospitalization, with pain at 0.5 weeks after admission for each score set to 100%. At 4 weeks post-admission, significant differences were observed among the three groups in the NRS and Abbey-J scores during exercise and in Doloplus-2 scores. No significant differences were observed at any other time points (one-way ANOVA at 1.0 week: F2, 98 = 2.175, p = 0.119; at 1.5 weeks: F2, 98 = 1.145, p = 0.322; at 2.0 weeks: F2, 93 = 0.0405, p = 0.322; at 2.5 weeks: F2, 79 = 0.657, p = 0.521; at 3.0 weeks: F2, 72 = 2.541, p = 0.0858; at 3.5 weeks: F2, 60 = 0.018, p = 0.982 (Fig 2); At 4.0 weeks, NRS: 2.72619 ± 2.169252, n = 22: Abbey-J: 0.588235 ± 0.901923, n = 20: Doloplus-2: 1.705882 ±1.611083, n = 21; one-way ANOVA followed by the bonferroni test: F2, 59 = 6.854, p = 0.00211) (Fig 2).

**Fig 2.**
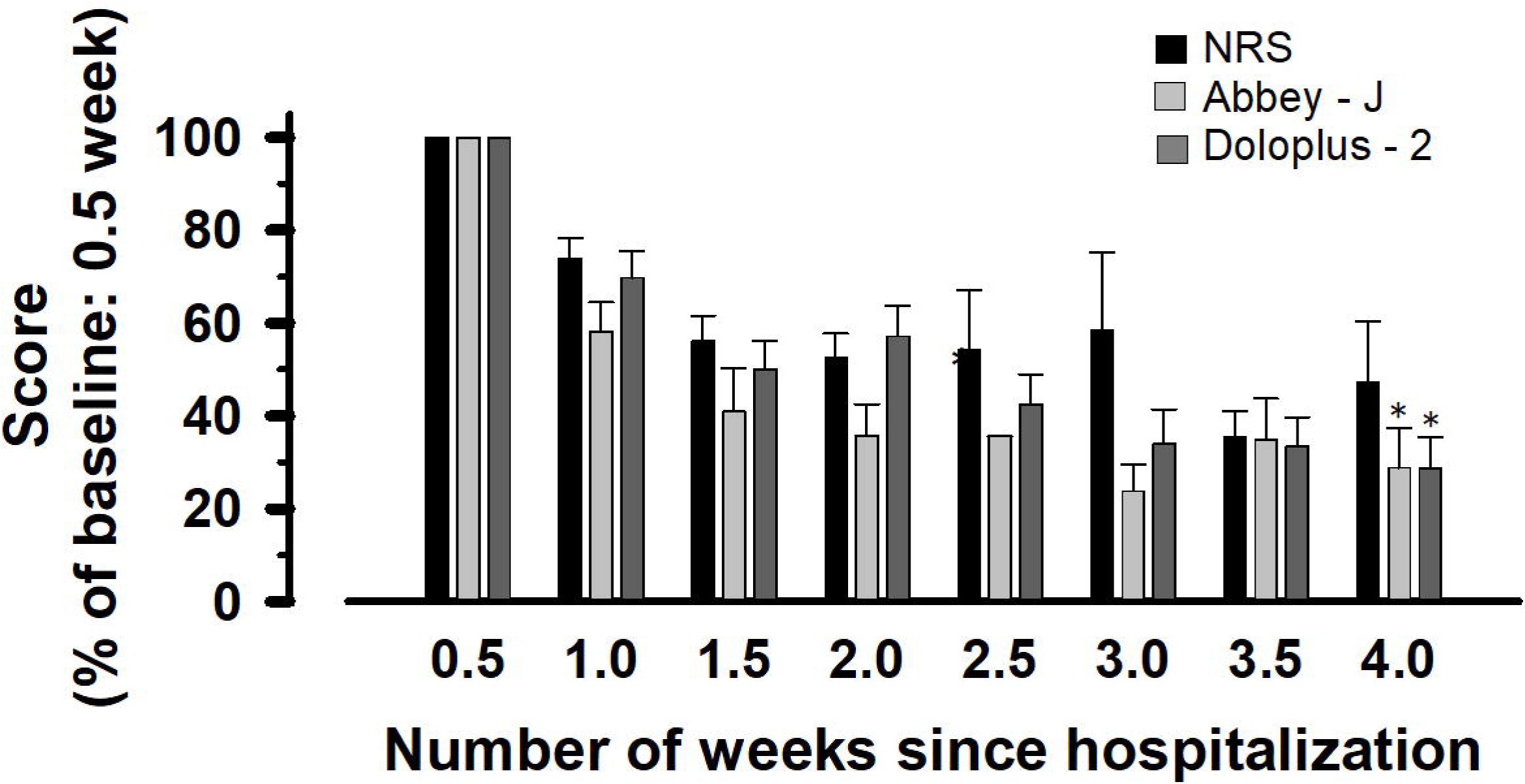
Changes in scores for each pain assessment tool of hospitalized patients with vertebral fractures. The pain score at admission is taken as 100. **p value < 0.01 is statistically significant. NRS, Numerical Rating Scale; Abbey-J, Japanese version of Abbey Pain Scale.

Although observational assessment tools may underestimate the pain compared with assessments made by patients, at 4 weeks post-admission, observational assessment tools and self-assessment tools did not significantly differ.

### Correlation between pain assessment tools

At 2 and 3 weeks post-admission, the NRS score was significantly positively correlated with both the Abbey-J and Doloplus-2 scores (Table 2).

**Table 2.**
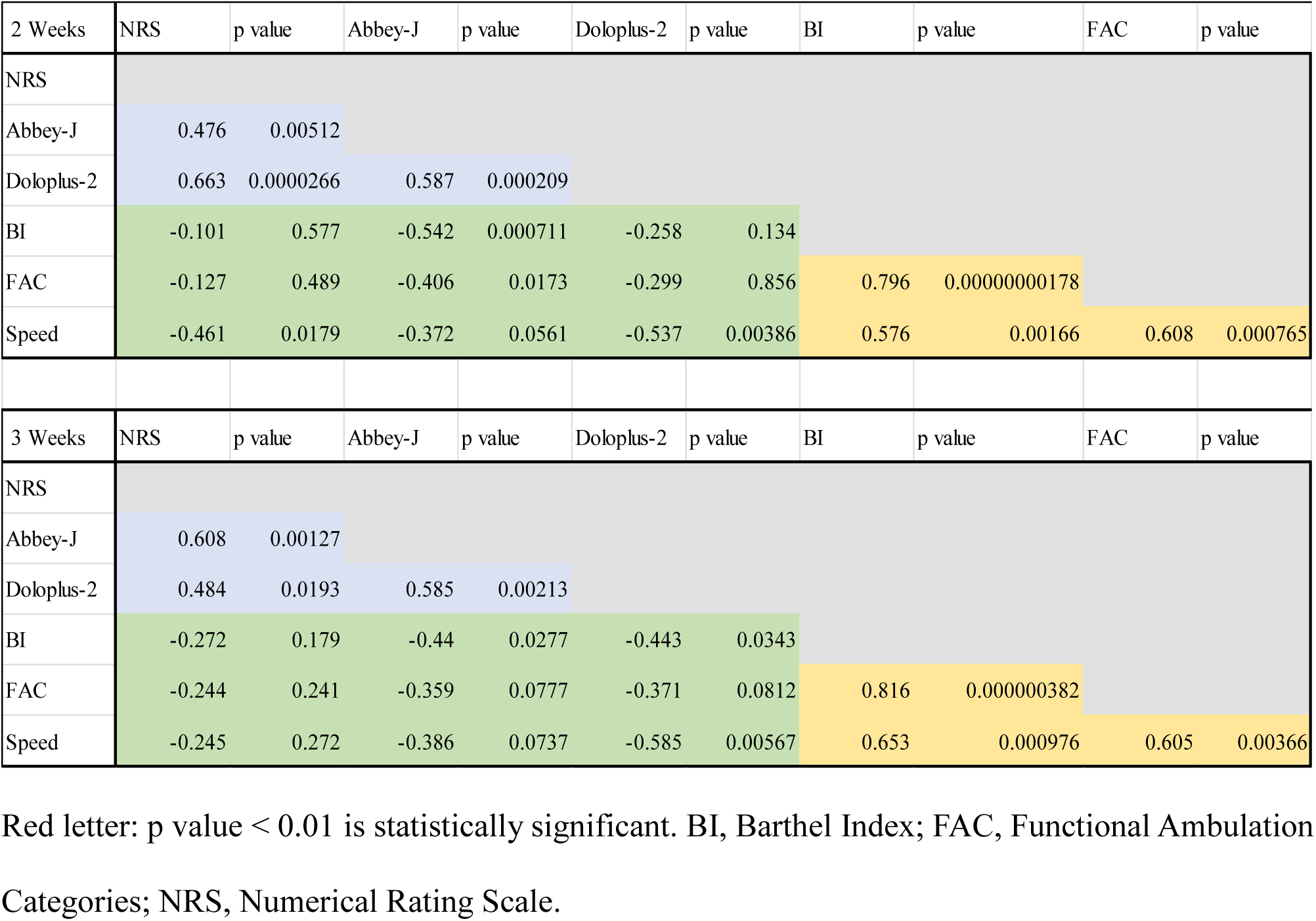
Correlation of each pain assessment tool with ADLs and ambulatory status (at weeks 2 and 3 of hospitalization)

### Changes in ADLs and ambulatory status over time

Ambulatory status evaluated using the FAC significantly improved over time after admission. ADLs evaluated using the BI and 10-m walking tests were not significantly different; however, there was an increasing trend (repeated-measures ANOVA followed by the bonferroni test; BI: F3,15 = 1.7476, p = 0.200282; at 1 weeks vs. at 2 weeks: p = 1, vs. at 3 weeks: p = 1, vs. at 4 weeks: p = 0.905; the FAC: F4,16 = 7.5909, p = 0.001255; at 0 weeks vs. at 1 weeks: p = 0.00024, vs. at 2 weeks: p = 0.00122, vs. at 3 weeks: p = 0.00132, vs. at 4 weeks: p = 0.05276; 10-m walking test: F3,3 = 10.414, p = 0.04285; at 1 week vs. at 2 weeks: p = 1, vs. at 3 weeks: p = 0.072, vs. at 4 weeks: p = 0.46) (Fig 3).

**Fig 3.**
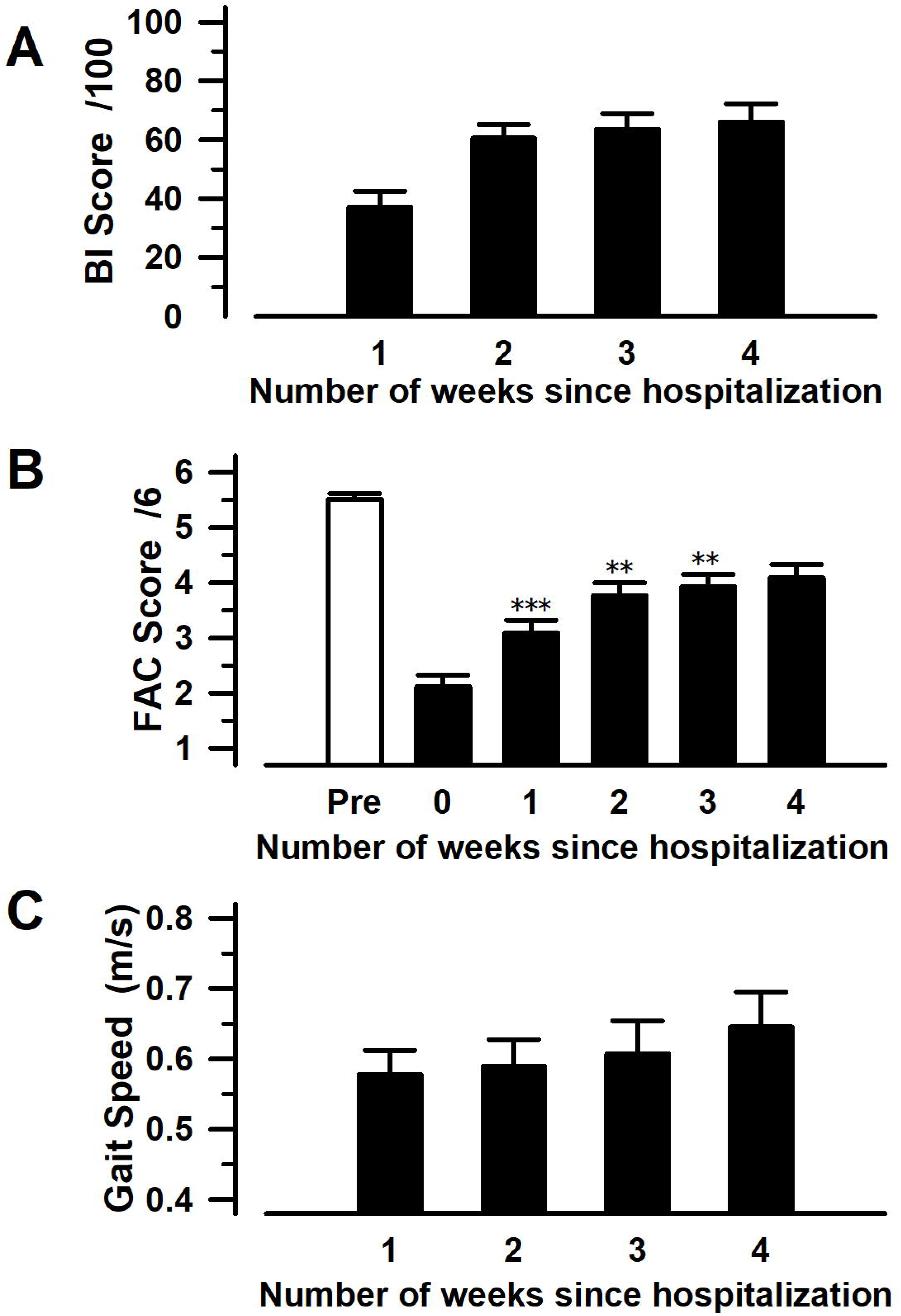
Changes in scores for ADLs and ambulatory status in hospitalized patients with vertebral fractures. (A) Barthel Index (BI); (B) Functional Ambulation Categories (FAC); (C) 10-m walking test.

### Correlation between ADLs and ambulatory status

At 2 and 3 weeks post-admission, the BI was significantly positively correlated with both the FAC and 10-m walking test, which are assessment tools for ambulatory ability (Table 2).

### Correlation of each pain assessment tool with ADLs and ambulatory status

At 2 and 3 weeks post-admission, each pain assessment tool showed a relatively negative correlation with the assessment tools for ambulatory status and ADLs (Table 2).

### Evaluation of scores assigned to items in each pain assessment tool according to MMSE scores

Each pain assessment tool was used to evaluate the pain of patients with vertebral fractures according to their degree of cognitive impairment. Of the six patients with an MMSE score of 17 or lower, two were able to assess their pain using the NRS. Pain was difficult to assess in many patients, indicating that the NRS is insufficient for pain assessment. The scores on the observational assessment tools were not significantly different between patients with an MMSE score of 17 or lower and those with an MMSE score of 18 or higher, and the scores significantly decreased within 3 weeks. However, only the Abbey-J assessment at 4 weeks showed a significant difference among the three MMSE groups (one-way ANOVA followed by the bonferroni test: Abbey-J (during movement) at 1 week: F2, 32 = 0.912, p = 0.412; at 2 weeks: F2, 32 = 0.78, p = 0.467; at 3 weeks: F2, 25 = 1.49, p = 0.245; at 4 weeks: F2, 19 = 7.088, p = 0.00502, MMSE 17 - 0 vs. MMSE 23 - 18: p = 1, vs. MMSE 30 - 24: p = 0.024, MMSE 30–24 vs. MMSE 23 - 18: p = 0.02) (Fig 4).

**Fig 4.**
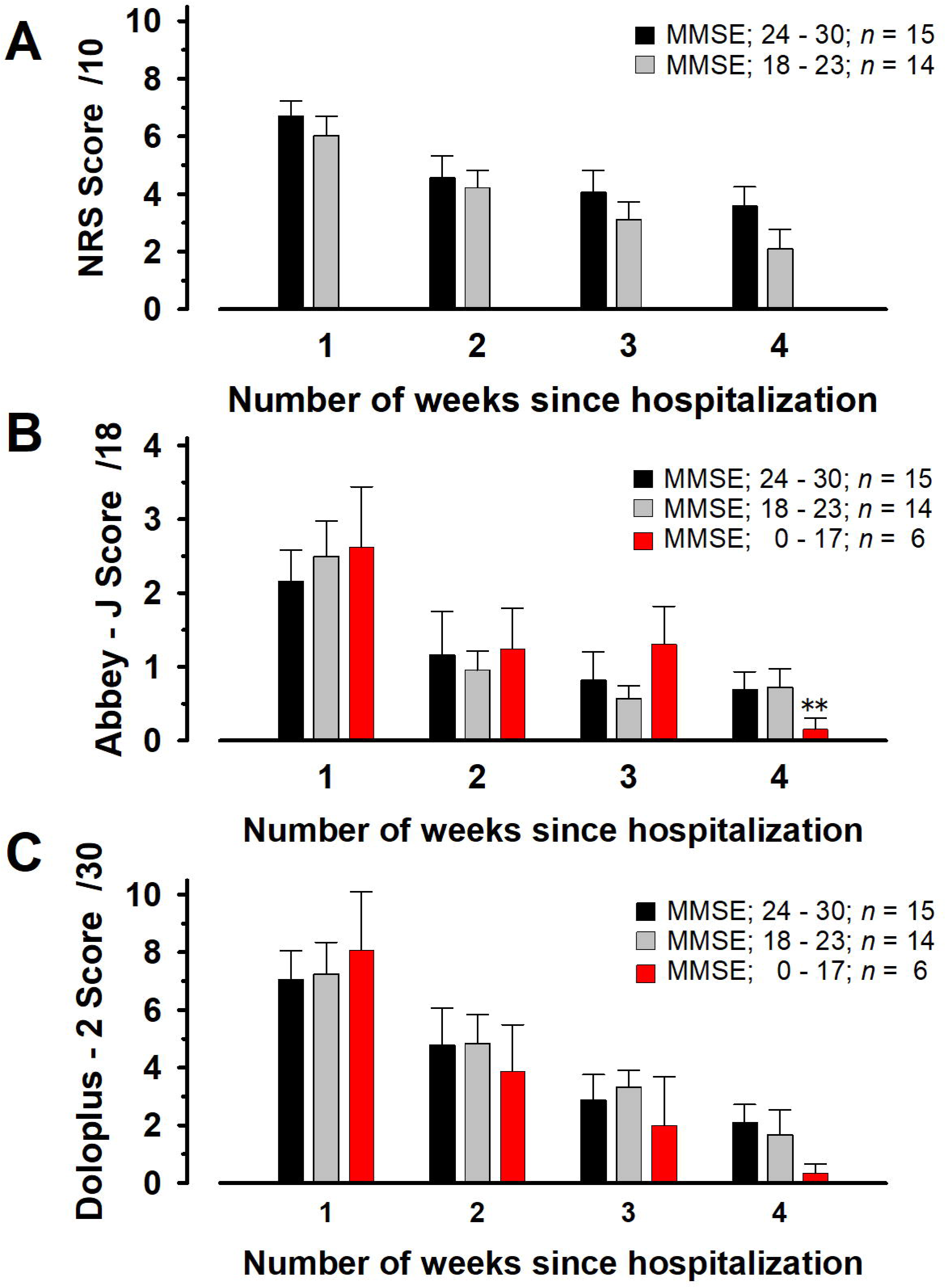
Pain assessment tool score changes for hospitalized patients with vertebral fractures per cognitive impairment degree. (A) Numerical Rating Scale (NRS); (B) Japanese version of Abbey Pain Scale (Abbey-J), (C) Doloplus-2. **p value < 0.01 is statistically significant. MMSE, Mini-Mental State Examination.

## Discussion

In this study, all NRS (a self-reported assessment tool), Abbey-J, and Doloplus-2 (observational assessment tools) scores decreased significantly over time after admission. The NRS score positively correlated with both the Abbey-J and Doloplus-2 scores. In addition, each pain assessment tool negatively correlated with ADLs and ambulatory status. Therefore, this study verified that observational assessment tools were valid for assessing pain in patients with acute-phase vertebral fractures. However, the Abbey-J score showed a significant difference only at 2.5 weeks post-admission. Since the Abbey-J is a relatively simple observational assessment tool, it may not be possible to adequately evaluate the transition from acute to chronic pain. Additionally, at 4 weeks post-admission, significant differences were observed among the three MMSE groups (normal, mild, and moderate or severe) in the NRS and Abbey-J scores during exercise, and in Doloplus-2 scores. Since observational assessment tools are objective, it is possible that patient pain was underestimated.

The NRS is an extremely simple self-reported pain assessment tool that is widely used in clinical settings and does not require writing instruments. The NRS can be used in patients with mild cognitive impairment, as defined by an MMSE score of 18 points or higher. However, for patients with an MMSE scores of 17 points or lower, NRS scores are reportedly difficult to record [12]. In contrast, the Abbey-J was developed as an observational tool to assess the pain intensity in individuals with dementia, and its use is also relatively simple [29]. In this study, the Abbey-J scores for low back pain associated with vertebral fractures were relatively low. One of the characteristics of this study was that pain was classified as mild in several patients. In addition, the pain scores on Doloplus-2 tended to decrease within a short period after admission. Pain scores evaluated with observational assessment tools may deviate from the actual severity of clinical symptoms (such as low back pain). However, the knowledge that pain decreases over time may be sufficient for pain assessment.

Of the patients examined with vertebral fractures, the self-reported assessment tool positively correlated with observational assessment tools. Considering that this study included older patients and those with cognitive impairment, our results demonstrated that the Abbey-J and Doloplus-2 were equivalent to the widely used NRS in the current clinical setting. These observational assessment tools may also be suitable to estimate the pain intensity associated with acute-phase orthopedic disorders that are common in older patients.

Furthermore, the results showing a positive correlation between BI and FAC in study weeks 2 and 3 indicated that ADLs can be determined by evaluating the ambulatory status in older patients with vertebral fractures, including those with cognitive impairment. In addition, the negative correlation of each pain assessment tool with ADLs and ambulatory status suggests that the observation of both ambulatory status and ADLs can contribute to pain assessment. Although the degree of correlation differed somewhat between the self-reported and observational assessment tools in this study, observational assessment tools may be used in the future, even for patients who have difficulty using the NRS to assess their pain. Since a self-reported pain assessment tool such as the NRS is a subjective approach, it is affected by different pain thresholds among individuals. Poor objectivity is one of the limitations of self-reported assessment tools. Thus, pain cannot be objectively assessed in patients for whom self-evaluation of pain is difficult, such as those with cognitive impairment. In this context, assessor objective approaches to scoring pain based on patient behavior appear to be easy to perform because the evaluation criteria had been determined [30, 31].

When the scores for each pain assessment tool were compared of patients with vertebral fractures according to the degree of cognitive impairment, pain tended to be assessed as more severe with the Abbey-J and Doloplus-2 than with the NSR in patients with cognitive impairment. It is difficult to determine which tool is superior or inferior based solely on these results. However, patients with cognitive impairment may not be able to understand, recognize, or express pain. Thus, their expression of pain may deviate from their behavior [32, 33]. The validity of the observational assessment tools demonstrated in this study may have implications in clinical settings because pain intensity can be determined based on behavior [34, 35]. The Abbey and Doloplus-2 have been reported to be appropriate for pain assessment in patients with moderate-to-severe dementia [29, 36]. However, evaluators may need to be proficient in observing patient conditions to assess nonverbal communication, such as facial expressions and behaviors, in greater detail [37]. Thus, the fact that there was a significant difference between the Abbey-J scores in patients with an MMSE score of ≤ 17 points and in patients with an MMSE score of ≥ 18 points at 4 weeks may suggest the need for proficient observational skills to be employed when assessing pain.

## Limitations

Observational assessment tools may underestimate pain in patients with mild-to-moderate pain and the scoring is prone to inter-evaluator differences. In observational assessments, pain other than low back pain may also be simultaneously and inadvertently assessed. Therefore, caution should be exercised when interpreting the results. In particular, it is necessary to monitor patients for the onset of comorbidities after admission as factors other than low back pain may substantially change the scores of the observational assessment tools.

## Conclusions

The self-reported NRS scores of patients who received conservative therapy for acute vertebral fractures at our hospital were significantly positively correlated with observational assessments using the Abbey-J or Doloplus-2. All pain assessment tools were significantly negatively correlated with ADLs and ambulatory status. The results of this study suggest that observational assessments using the Abbey-J or Doloplus-2 could be used to estimate pain, even in patients with cognitive impairment where self-reported assessments with the NRS are difficult to perform.

## Data Availability

All data produced in the present study are available upon reasonable request to the authors

## Acknowledgments

We thank the medical staff of Moji Medical Center, Kitakyushu, Japan, for their assistance. We would like to thank Editage for English language editing.

## Supporting information captions

**S1 Table. Patient characteristics.**

**S2 Table. Correlation of each pain assessment tool with ADLs and ambulatory status (weeks 2 and 3 of hospitalization).** Red letter, p value < 0.01 is statistically significant. ADLs, activities of daily living.

**S1 Fig. Changes in scores for each pain assessment tool in hospitalized patients with vertebral fractures.** (A) Numerical Rating Scale (NRS); (B) Japanese version of Abbey Pain Scale (Abbey-J) (C) Doloplus-2. **p value < 0.01 and ***p value < 0.01 is statistically significant.

**S2 Fig. Changes in scores for each pain assessment tool in hospitalized patients with vertebral fractures (pain score at admission is taken as 100).** **p value < 0.01 is statistically significant.

**S3 Fig. Changes in scores for ADLs and ambulatory status in hospitalized patients with vertebral fractures.** (A) Barthel Index (BI); (B) Functional Ambulation Categories (FAC); (C) 10-m walking test; ADLs, activities of daily living

**S4 Fig. Changes in scores for each pain assessment tool of hospitalized patients with vertebral fractures according to the degree of cognitive impairment.** (A) Numerical Rating Scale (NRS); (B) Japanese version of Abbey Pain Scale (Abbey-J); (C) Doloplus-2. **p value < 0.01 is statistically significant.

